# Hospitalisation rates differed by city district and ethnicity during the first wave of COVID-19 in Amsterdam, the Netherlands

**DOI:** 10.1101/2021.03.15.21253597

**Authors:** Liza Coyer, Elke Wynberg, Marcel Buster, Camiel Wijffels, Maria Prins, Anja Schreijer, Yvonne T.H.P van Duijnhoven, Alje van Dam, Mariken van der Lubben, Tjalling Leenstra

**Affiliations:** Department of Infectious Diseases, Public Health Service of Amsterdam, Amsterdam, the Netherlands; Department of Infectious Diseases, Amsterdam Infection & Immunity (AII), Amsterdam UMC, University of Amsterdam, Amsterdam, the Netherlands; Department of Epidemiology, Health Promotion and Care Innovation, Public Health Service of Amsterdam, Amsterdam, the Netherlands

**Author notes:** Corresponding author: Liza Coyer, Public Health Service of Amsterdam Nieuwe Achtergracht 100, 1018 WT Amsterdam, the Netherlands. Contributed equally (Liza Coyer, Elke Wynberg).

**Keywords:** SARS-CoV-2, COVID-19, hospitalisation, ethnicity, geographical, socio-economic status

## Abstract

**Background:** It is important to gain insight into the burden of COVID-19 at city district level to develop targeted prevention strategies. We examined COVID-19 related hospitalisations by city district and migration background in the municipality of Amsterdam, the Netherlands.

**Methods:** We used surveillance data on all PCR-confirmed SARS-CoV-2 hospitalisations in Amsterdam until 31 May 2020, matched to municipal registration data on migration background. We calculated directly standardised (age, sex) rates (DSR) of hospitalisations, as a proxy of COVID-19 burden, per 100,000 population by city district and migration background. We calculated standardised rate differences (RD) and rate ratios (RR) to compare hospitalisations between city districts of varying socio-economic and health status and between migration backgrounds. We evaluated the effects of city district and migration background on hospitalisation after adjusting for age and sex using Poisson regression.

**Results:** Between 29 February and 31 May 2020, 2326 cases (median age 57 years [IQR=37-74]) were notified in Amsterdam, of which 596 (25.6%) hospitalisations and 287 (12.3%) deaths. 526/596 (88.2%) hospitalisations could be matched to the registration database. DSR were higher in individuals living in peripheral (South-East/New-West/North) city districts with lower economic and health status, compared to central districts (Centre/West/South/East) (RD=36.87,95%CI=25.79-47.96;RR=1.82,95%CI=1.65-1.99), and among individuals with a non-Western migration background compared to ethnic-Dutch individuals (RD=57.05,95%CI=43.34-70.75; RR=2.36,95%CI=2.17-2.54). City district and migration background were independently associated with hospitalisation.

**Conclusion:** City districts with lower economic and health status and those with a non-Western migration background had the highest burden of COVID-19 during the first wave of COVID-19 in Amsterdam.

## INTRODUCTION

The first cases of coronavirus disease 2019 (COVID-19) were reported at the end of 2019 in Wuhan, China. Severe acute respiratory syndrome coronavirus 2 (SARS-CoV-2), the virus causing COVID-19, has since rapidly spread across the globe. The World Health Organization (WHO) consequently declared COVID-19 a global epidemic on 11 March 2020. As of 29 January 2021, over 100 million cases and more than 2 million COVID-19 related deaths had been reported worldwide(1).

In the Netherlands, COVID-19 was declared a Group A notifiable disease under the Public Health legal act on 28^th^ January 2020(2). This required clinicians and laboratories to immediately notify the regional public health service (PHS) every suspected and/or confirmed case of COVID-19. At the same time, sporadic introductions following imported cases from Asia were identified in France, Germany and Northern Italy(3). The first confirmed case of COVID-19 in Tilburg on 27 February 2020 had an epidemiological link to Northern Italy. Initial sporadic clusters led to a subsequent nationwide spread of the virus, causing significant morbidity and mortality, including in the capital city of Amsterdam. From 12-15 March, the Dutch government initiated a series of restrictions as part of a so-called “intelligent lockdown”, including cancellation of events with 100+ attendees, working from home wherever possible, and the closure of public facilities(4). These measures were gradually lifted from 11 May onwards with the reopening of schools and contact professions.

The regional PHS of Amsterdam acts upon notifications of infectious diseases including COVID-19 for the wider Amsterdam-Amstelland area, consisting of 6 municipalities including Amsterdam, with a total regional population of about 1,06 million. Anecdotal reports from hospital staff in April 2020 suggested that a disproportionate number of patients of ethnic minority background had been admitted to hospitals and intensive care units (ICU) in Amsterdam. This echoed reports from the United Kingdom(5) (UK) and United States of America(6) (USA) at the time, which suggested that age- and sex-standardised mortality rates were highest among socio-economically deprived groups and ethnic minority groups; findings that were later supported by a systematic review(7). Early analyses also demonstrated that individuals with a migration background in the Netherlands had higher rates of excess mortality (i.e. higher than would be expected in ‘normal’ conditions) compared to individuals of Dutch origin, in particular in those with a non-Western migration background(8). Amsterdam is an increasingly ethnically diverse city with more than half the population having a migration background(9). Disparities in the distribution of communicable and non-communicable diseases have previously been demonstrated in the city between individuals with and without a migration background(10), even when matched for age- and socio-economic status (SES)(11). In addition, health inequalities have been reported between the peripheral city districts of Amsterdam with lower average income (12) and the central, higher-income city districts of the Centre, West and East districts, with poorer health outcomes being reported in peripheral districts(13). Investigating whether such disparities also exist for COVID-19 burden is crucial to developing targeted prevention strategies.

We therefore aimed to examine differences in burden of COVID-19 hospitalisations between city districts and ethnic groups in the city of Amsterdam, using routinely collected surveillance data until the end of May 2020.

## METHODS

### COVID-19 case definition and test strategy

Up to and including 11^th^ March 2020, SARS-CoV-2 testing in the Netherlands was conducted only in individuals who fulfilled a strict case definition: (i) having an epidemiological link to a confirmed case and/or returning from a high-risk region with widespread transmission within 14 days prior to the onset of symptoms, and (ii) the presence of fever with at least one of the following symptoms: coughing, shortness of breath (dyspnoea). Testing was mainly carried out by a home testing team of the PHS, by general practitioners and in hospitals.

With the start of the intelligent lockdown from 12-15 March onwards, testing no longer required a confirmed epidemiological link or travel history, but instead focussed on mitigating the impact on frontline healthcare services and protecting vulnerable groups. Healthcare workers, residents of long-term care facilities and individuals at high risk of severe disease with COVID-like symptoms were prioritised. For patients, testing was carried out in the hospital setting (including purpose-built COVID-19 triage and testing tents), by family physicians, and by the PHS’s home testing team, who also performed testing in long-term care facilities. Healthcare workers could be tested at a special test location at the PHS site itself or at work (in hospitals) and were prioritised in situations where a staffing shortage was at stake.

From 11 May 2020, testing at the PHS site was also made available to teachers and those working in other contact professions with COVID-like symptoms. Subsequently from 1 June 2020 testing became available for all individuals with at least 24 hours of COVID-like symptoms. Because of the restrictive test policy until 1 June 2020, we used hospitalisations as a marker of epidemic progression in this study.

### Data collection and linkage

After 28^th^ January 2020, all persons with a positive PCR test for SARS-CoV-2 were required to be notified to the PHS, which subsequently notified the National Institute for Public Health and the Environment (RIVM). In the wider region of Amsterdam-Amstelland, all positive cases were notified to the regional PHS of Amsterdam. For this study, we retrieved surveillance data from all notified cases, hospitalisations and deaths residing in the municipality of Amsterdam between 29 February (date of first confirmed case) and 31 May 2020. These data included information on age and sex of the case, and whether the case had worked as a health care worker or was a resident in a long-term care facility. We matched surveillance data to registration data from the municipality records of the City of Amsterdam (BRP) to retrieve postal code and the country of birth of the case and their parents.

### Determination of city district and migration background

We determined city district based on postal code of current residence(14). We assessed migration background and generation based on the country of birth of individuals and their parents(15). Dutch ethnic origin, i.e. having no migration background, was defined as having parents who were both born in the Netherlands. An individual was considered of non-Dutch ethnicity if he/she was born abroad and had at least one parent who was born abroad (first generation) or he/she was born in the Netherlands and both parents were born abroad (second generation)(16). For second generation migrants of whom both parents were born abroad, the mother’s country of birth was leading in defining migration background. Individuals of non-Dutch ethnicity were further classified into having a non-Western migration background (from African, Latin-American or Asian countries, or Turkey, excluding Indonesia and Japan) or Western migration background (from North-American, European or Oceanian countries, or Indonesia or Japan, excluding Turkey) according to the definition of the Dutch Central Bureau of Statistics(16).

### Statistical analysis

We evaluated the number of cases, hospitalisations and deaths, overall and over time (notification date and symptom onset). In the epidemiological curve by self-reported date of symptom onset, missing dates were imputed, assuming that all those who were tested had symptoms. First, we created a distribution of time between symptom onset and case notification for those with a known date of symptom onset. Second, we randomly sampled from this distribution to estimate the date of symptom onset if this was missing.

We calculated crude and directly standardised rates (DSR) of hospitalisations per 100,000 population, overall and by city district and migration background. Rates were standardised for age (≤14, 15-29, 29-44, 45-59, 60-74, ≥75 years) and sex (female, male). 95% confidence intervals (CI) were calculated using the gamma method(17,18). We computed rate differences (RD) and rate ratios (RR) to compare DSR of hospitalisation between (i) city districts, (ii) migration backgrounds (first and second generation migrants combined) and (iii) a combination of both variables, i.e. six strata of city districts (dichotomized into the central districts with higher average incomes (Central/West/South/East) and peripheral districts with lower average incomes (Southeast/North/New-West), based on 2018 income per capita per city district(12)) and migration background (none [ethnic-Dutch], Western, non-Western). First and second generation were combined as this was highly correlated with age and a stratified analysis would result in few outcomes in specific age-stratums. Since elderly persons with a migration background are less likely to engage with home care or nursing homes (20), we hypothesized that they would be more likely to be hospitalised upon a deterioration in health than ethnic-Dutch elderly, who may be more likely to receive palliative care in the community care setting. To explore this possible bias, we conducted an additional analysis of DSR of hospitalisation by migration background, stratified by age <60 and ≥60 years.

We evaluated the individual effects of city district (peripheral, central), migration background, sex and age in a Poisson regression model, using the log of the population size per district/background/sex/age stratum as an offset. In this model, we added an interaction term between city district and migration background which was tested for significance using a likelihood ratio rest.

We assumed statistical significance at a *P*-value<0·05. We used the dsr package in R(18) to calculate DSR, RD and RR. All analyses were performed in R (version 3.6.3, Vienna, Austria).

## RESULTS

Between 29 February 2020 and 31 May 2020, 2326 COVID-19 cases were notified in Amsterdam, of which 596 (25.6%) hospitalisations and 287 (12.3%) deaths. The number of new cases peaked mid-March, shown by date of symptom onset in Figure 1 and by notification date in Supplementary Figure S1. Characteristics of COVID-19 cases by hospitalisation status are presented in Table 1. Median age of hospitalised cases was higher (64 [IQR 51-73]) than cases for which hospitalisation status was non-hospitalised or unknown (54 [IQR 34-74]). The majority of hospitalised cases were male (361/596 [60.6%]), while most cases overall were female (1346 [57.9%]). 814 (35.0%) of all notifications were registered as health care workers, of whom 314 (38.6%) worked in a long-term care facility.

**Table 1.** Characteristics of COVID-19 cases and hospitalisations in Amsterdam, the Netherlands, 29 February-31 May 2020

| Characteristic | Total<br>(N=2326) |  | Hospitalised<br>(n=596) |  | Not hospitalised/<br>unknown (n=1730) |  |
| --- | --- | --- | --- | --- | --- | --- |
|  | n | % | n | % | n | % |
| **Age in years, median [IQR]** | 57 | [37-74] | 64 | [51-73] | 54 | [34-74] |
| **Sex** |  |  |  |  |  |  |
| Female | 1346 | 57.9 | 230 | 38.6 | 1116 | 64.5 |
| Male | 965 | 41.5 | 361 | 60.6 | 604 | 34.9 |
| Unknown | 15 | 0.6 | 5 | 0.8 | 10 | 0.6 |
| **Health care worker (HCW)** |  |  |  |  |  |  |
| No | 1108 | 47.6 | 383 | 64.3 | 725 | 41.9 |
| Yes | 814 | 35.0 | 47 | 7.9 | 767 | 44.3 |
| HCW not in a long-term care facility/unknown location | 500 | 61.4 | 36 | 76.6 | 464 | 60.5 |
| HCW in a long-term care facility | 314 | 38.6 | 11 | 23.4 | 303 | 39.5 |
| Unknown | 404 | 17.3 | 166 | 27.8 | 238 | 13.8 |
| **Resident of a long-term care facility** |  |  |  |  |  |  |
| No/unknown | 1867 | 80.3 | 579 | 97.2 | 1299 | 74.5 |
| Yes | 459 | 19.7 | 17 | 2.9 | 445 | 25.5 |
| **City district** |  |  |  |  |  |  |
| Centre | 162 | 7.0 | 37 | 6.2 | 141 | 7.0 |
| New-West | 433 | 18.6 | 130 | 21.8 | 360 | 18.0 |
| North | 268 | 11.5 | 82 | 13.8 | 239 | 12.0 |
| East | 377 | 16.2 | 62 | 10.4 | 330 | 16.5 |
| West | 341 | 14.7 | 86 | 14.4 | 293 | 14.6 |
| South | 350 | 15.1 | 77 | 12.9 | 312 | 15.6 |
| South-East | 355 | 15.3 | 113 | 19.0 | 320 | 16.0 |
| Unknown | 40 | 1.7 | 9 | 1.5 | 7 | 0.4 |
| **Died** |  |  |  |  |  |  |
| No/unknown | 2039 | 87.7 | 496 | 83.2 | 1543 | 89.2 |
| Yes | 287 | 12.3 | 100 | 16.8 | 187 | 10.8 |

Abbreviations: IQR, interquartile range

**Figure 1.**
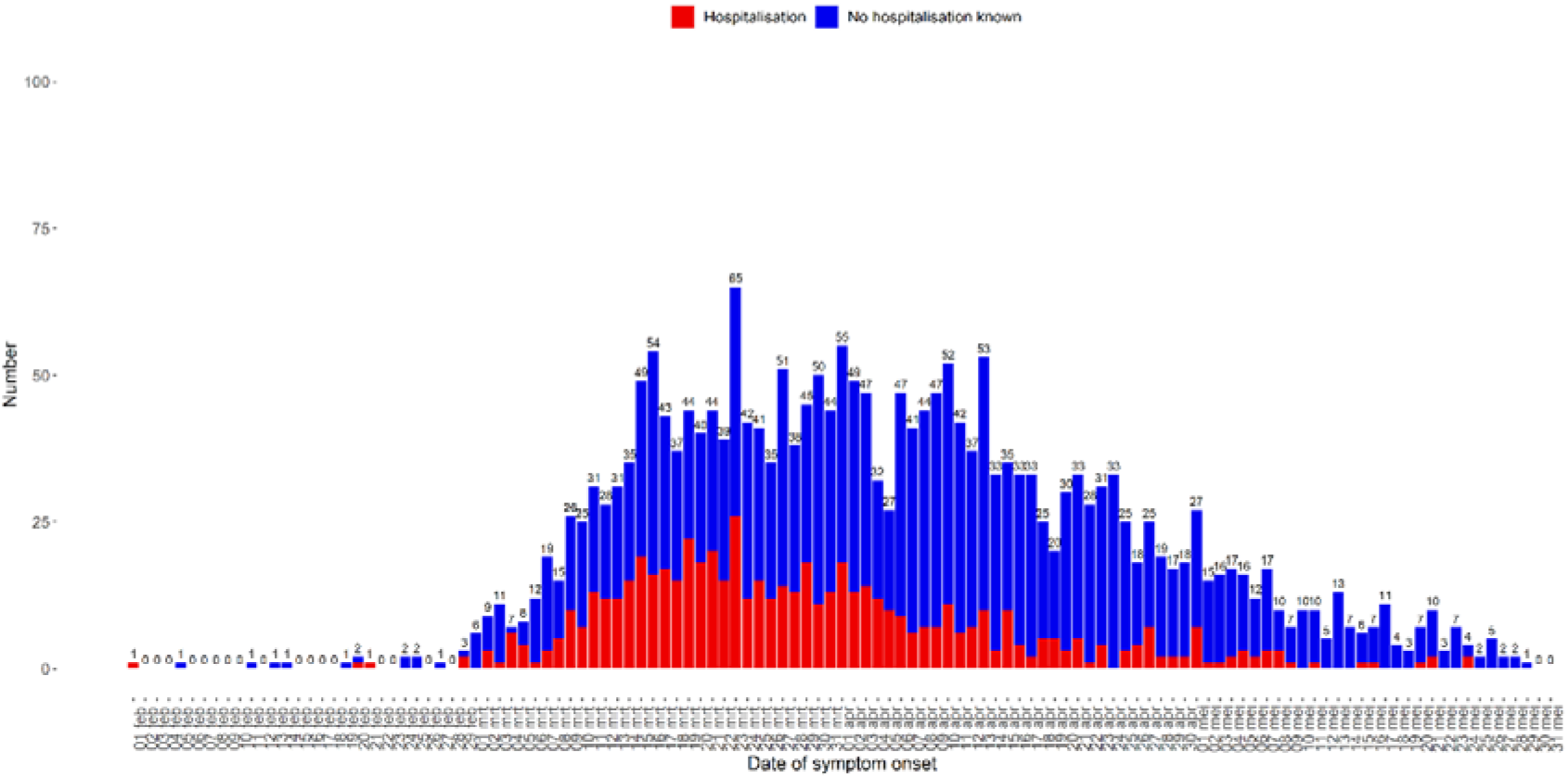
COVID-19 notifications in Amsterdam between 29 February and 31 May 2020 by (imputed) date of symptom onset, stratified by hospitalisation status

Of the 2326 cases, 2002 (86.1%) could be matched to the registration database, including 526/596 hospitalisations (88.2%) and 235/287 (81.8%) deaths (Supplementary Figure S2, Supplementary Table S1). Table 2 shows hospitalisation DSR per 100,000 population by city district and migration background in the matched population. Hospitalisation DSR were almost two-fold higher in individuals living in peripheral city districts compared to central districts (RR=1.82, 95%CI=1.65-1.99), with 36.87 additional hospitalisations per 100.000 population (RD=36.87, 95%CI=25.79-47.96). The difference between peripheral and central city districts became increasingly apparent over time (Figure S3). Further stratification by individual city district is presented in Supplementary Table S2.

**Table 2.** Hospitalisation rates by city district and migration among those linked to the registration database, Amsterdam, the Netherlands, 29 February – 31 May 2020

|  | Hospital admissions | Population<sup>a</sup> | Crude rate per 100,000 population (95% CI) | Standardised rate per 100,000 population<sup>b</sup> (95% CI) | Standardised rate difference (95% CI) | Standardised rate ratio (95% CI) |
| --- | --- | --- | --- | --- | --- | --- |
| **Total<sup>c</sup>** | 523 | 873,055 | 59.90<br>(54.88-65.27) |  |  |  |
| **City district** |  |  |  |  |  |  |
| Central | 233 | 523,536 | 44.51<br>(38.97-50.6) | 44.99<br>(39.40-51.16) | Ref. | Ref. |
| Peripheral | 289 | 349,519 | 82.69<br>(73.43-92.79) | 81.87<br>(72.68-91.89) | 36.87<br>(25.79-47.96) | 1.82<br>(1.65-1.99) |
| **Migration background** |  |  |  |  |  |  |
| None (ethnic-Dutch) | 204 | 386,521 | 52.78<br>(45.78-60.54) | 42.04<br>(36.38-48.32) | Ref. | Ref. |
| Non-Western | 262 | 316,720 | 82.72<br>(73.01-93.37) | 99.08<br>(87.08-112.28) | 57.05<br>(43.34-70.75) | 2.36<br>(2.17-2.54) |
| Western | 57 | 169,814 | 33.57<br>(25.42-43.49) | 41.06<br>(30.96-53.4) | -0.98<br>(-13.28-11.32) | 0.98<br>(0.68-1.27) |

<sup>a</sup> Population on 1 April 2020
<sup>b</sup> Standardised for age (in 15-year groups) and gender, using the total population of Amsterdam as the standard population
<sup>c</sup> 3 matched cases had missing data

Hospitalisation DSR were more than two-fold higher in individuals with a non-Western migration background compared to ethnic-Dutch individuals (RR=2.36, 95%CI=2.17-2.54), with 57 additional hospitalisations per 100.000 population (RD=57.05, 95%CI=43.34-70.75). No statistically significant difference was observed between individuals with a Western migration background and ethnic-Dutch individuals. Further stratification by specific large non-Western ethnic groups living in Amsterdam (Dutch Antillean, Moroccan, Surinamese, Turkish and Ghanaian) is presented in Supplementary Table S3. Compared to the ethnic-Dutch population, the hospitalisation RR were highest among individuals of Ghanaian (RR=4.25, 95%CI=3.31-5.19; RD=136.75, 95%CI=-29.83-303.33) and Turkish ethnic origin (RR=3.08, 95%CI=2.72-3.43; RD=87.31, 95%CI=44.51-130.11). In our additional age-stratified analysis, we found that individuals <60 years with a non-Western migration background had a three-fold higher DSR compared to ethnic-Dutch individuals <60 years (RR=3.18, 95%CI=2.85-3.52), while the DSR was reduced but remained almost two-fold higher in individuals ≥60 years (RR=1.83, 95% CI: 1.57-2.09) (Supplementary Tables S4 and S5).

After stratifying by city district, the DSR was highest in non-Western residents of peripheral districts (Supplementary Figure S4): three times greater than in ethnic-Dutch residents of central districts RR=3.13, 95%CI=2.88-3.38; RD=76.64, 95%CI=57.38-95.89) (Figure 2).

**Figure 2.**
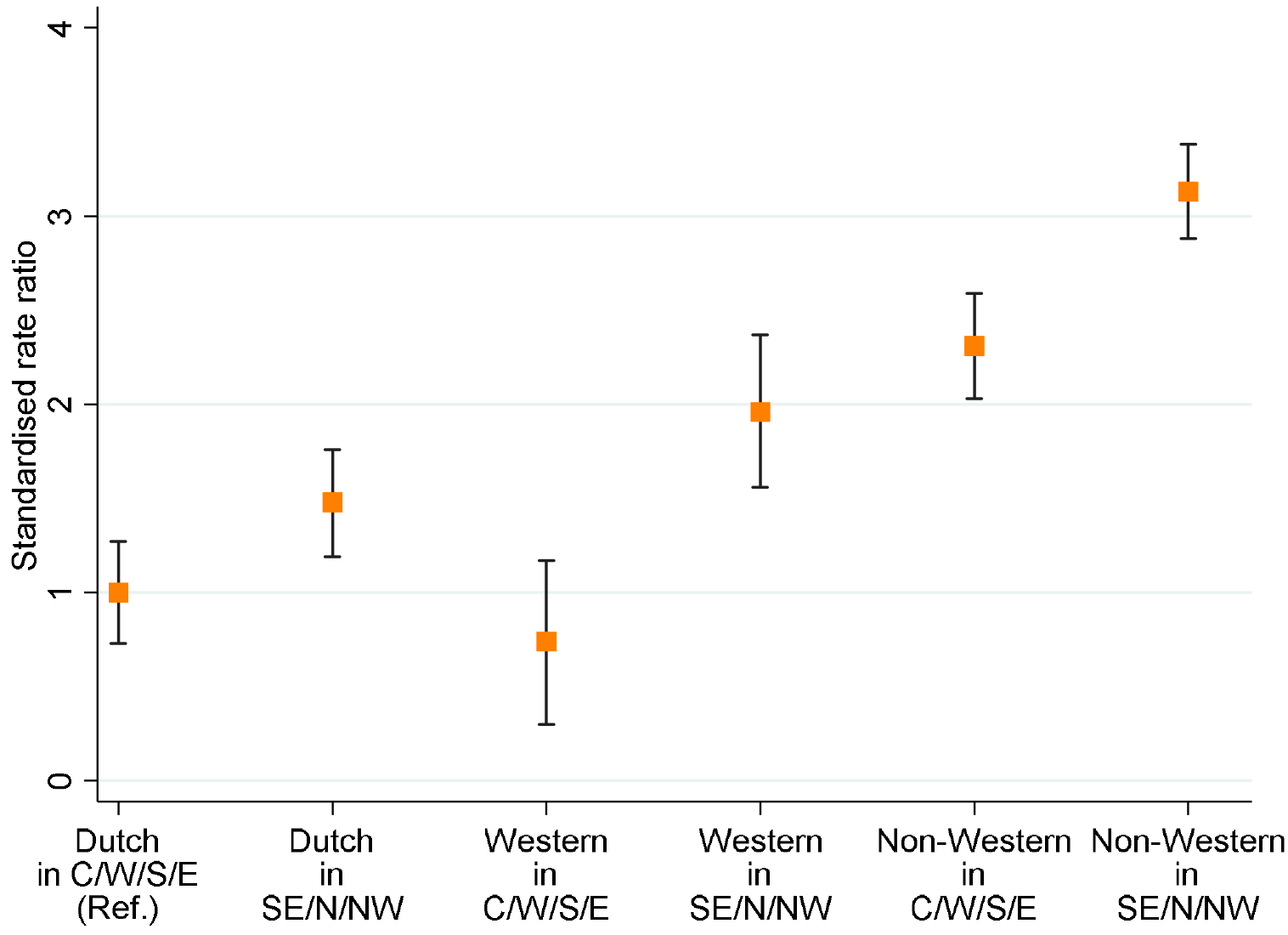
Standardised rate ratios and corresponding 95% CI comparing hospitalisation rates per 100,000 population by migration background and city district in the municipality of Amsterdam until 31 May 2020

In multivariable Poisson regression, non-Western migration background, living in a peripheral city district, male sex and older age were associated with hospitalisation(Table 3). No statistically significant interaction was found between migration background and city district (Supplementary Table S6) (*P=*0.08).

**Table 3.** Characteristics associated with COVID-19 related hospitalization, Amsterdam, the Netherlands, February-June 2020

| Characteristic | aRR (95% CI) | P-value |
| --- | --- | --- |
| **Migration background** |  |  |
| None (ethnic-Dutch) | 1 |  |
| Western | 0.97 (0.71-1.29) | 0.83 |
| Non-Western | 2.20 (1.82-2.67) | <0.001 |
| **City district** |  |  |
| Central (C/W/S/E) | 1 |  |
| Peripheral (SE/N/NW) | 1.53 (1.28-1.82) | <0.001 |
| **Sex** |  |  |
| Male | 1 |  |
| Female | 0.59 (0.49-0.70) | <0.001 |
| **Age** |  |  |
| <45 years | 1 |  |
| 45-59 years | 4.95 (3.73-6.6) | <0.001 |
| 60-74 years | 13.52 (10.47-17.63) | <0.001 |
| ≥75 years | 23.84 (17.92-31.88) | <0.001 |

Abbreviations: aRR, adjusted rate ratio; CI, confidence interval Estimates were obtained from a multivariable Poisson regression model.

## DISCUSSION

We identified geographical and ethnic disparities in the burden of COVID-19 in Amsterdam during the first wave of COVID-19. Specifically, people living in peripheral, lower-income city districts (New-West, North or South-East) with a non-Western migration background had three-fold higher hospitalisation rates compared to ethnic-Dutch individuals living central, higher-income city districts (Centre, West, South or East).

Previous research has demonstrated similar health disparities between city districts in Amsterdam. A quadrennially repeating comprehensive health survey has consistently shown that residents of peripheral city districts are more likely to suffer from comorbidities and be overweight or obese than residents of central districts(13). As chronic lifestyle conditions such as type 2 diabetes mellitus and cardio-vascular disease have been shown to increase the risk of developing severe COVID-19 symptoms(21), this suggests that the higher rate of COVID-19 hospitalisations seen in these city districts might be partially explained by a higher prevalence of chronic comorbidities. In addition, the peripheral districts have been shown to have a higher percentage of vulnerable residents(13), based on a vulnerability score that encompasses income, job security and healthcare expenditure. The interplay between SES and risk of both SARS-CoV-2 infection and severe disease has been previously documented(22). For example, individuals with a lower SES may be more likely to work as essential frontline workers – including healthcare workers and those working in sectors such as cleaning, construction, and transport(26) – with a reduced opportunity to work from home(25). Lower SES may also lead to a greater reliance on public transport, larger multi-generational households and smaller house size per person(26), all of which may increase the risk of exposure to infection(23). We therefore postulate that the greater burden of COVID-19 hospitalisation seen in these districts is due to a complex interaction between different factors. Indeed, when restricting to individuals <60 years of age in our analyses, the hospitalisation RR in those with a non-Western migration background compared to the ethnic-Dutch group remained and was even accentuated. This suggests that the differences in hospitalisation rates cannot be fully explained by an increased tendency of non-Western migrants, compared to ethnic-Dutch elderly, to be treated in hospital instead of receiving palliative care in the community setting or in nursing homes. The higher RR in younger age groups might imply that increased exposure secondary to having a public-facing occupation or higher levels of other causes of increased exposure among those with a non-Western migration background may have played an important role. Further elucidating the causal factors underlying SES disparities in COVID-19 burden requires further research.

In addition, our analysis demonstrated that individuals with a non-Western migration background, including groups with a Moroccan, Turkish, Surinamese and Ghanaian background, had a higher COVID-19 burden, even when stratifying by city district. This is in line with previous findings from the UK and USA and, more recently, other high-income countries such as Norway(27). In the UK, the risk of death among those with a COVID-19 diagnosis was twice as high for people of Bangladeshi ethnicity compared to people of White ethnicity, after adjusting for sex, age, deprivation and region(22). Additional studies demonstrated that, even when adjusting for age, sex, comorbidities and several SES-related determinants (though notably not public-facing occupation), hospitalisation and ICU admission rates among Black and Asian Minority Ethnic individuals were higher compared to ethnically White individuals(28), suggesting that ethnicity plays an independent role in explaining these disparities. In the US, age-adjusted hospitalisation rates were 5.3 and 4.7 times higher in American Indian or Alaska Native and Black or African American persons respectively, compared to non-Hispanic white persons; although this analysis was not adjusted for social inequalities(6). Whilst these findings consistently demonstrate that the burden of COVID-19 varies by ethnicity, the contribution of underlying cultural norms, health literacy, and differences in health-seeking behaviour has yet to be revealed.

Considering our findings substantiate reports of increased COVID-19 burden among ethnic minority groups in other countries, it is concerning that information on ethnicity is often not collected in routine surveillance systems in many countries(29). This results in these important differences in disease burden being concealed within the data, hindering the ability to set up targeted initiatives to reach particularly vulnerable populations. As stated by Tai et al.(30), further research on the interplay between inherent social inequalities and ethnicity is clearly required to ensure optimal surveillance of the impact of COVID-19 on vulnerable groups, to disentangle whether the increased burden is the result of a higher infection rate, the severity of disease manifestation, or both, and to inform and develop appropriate and accessible prevention strategies.

An important limitation of our study is that the surveillance data paint an incomplete picture of the first wave of the outbreak, as cases were underreported due to selective testing and data collection was limited. We used the hospitalisation rate per 100,000 population as a marker of outbreak progression, but this limits the distinction that can be made between risk of infection and risk of severe disease requiring hospital admission. In addition, hospitalisations and deaths among already notified cases may also have been underreported, and we were unable to match all notifications to the municipal register. Furthermore, absence of key individual socio-demographic, socio-economic, and clinical characteristics limits the inferences that can be made about causal factors on an individual patient level. For example, we used city district as an imperfect proxy for SES, but SES at the community or individual level within each city district may have been different. By further stratifying groups by migration background and complementing this with qualitative research (for example, through community focus groups) more insight can be gained into which community-specific targeted prevention strategies may help minimise the disproportionate distribution of COVID-19 in Amsterdam.

Our study is the first in the Netherlands to link surveillance data with registration data on migration background to demonstrate the unequal distribution of the burden of COVID-19 within the city of Amsterdam. We show that substantial differences in COVID-19 hospitalisation rates exist between city districts and ethnic groups in Amsterdam. Our findings corroborate reports from other high-income countries to suggest that public health efforts worldwide must be focussed on mitigating further impact of COVID-19 upon communities at highest risk of both infection and serious disease. Action must be taken to strengthen targeted prevention strategies which address the needs of affected communities.

## Data Availability

Information can be obtained from the corresponding author, Liza Coyer.

## Author contributions

LC, MB, and CW conducted data analyses, which were overseen by TL. LC, EW, MB, CW, MP, YvD and TL contributed to the interpretation of results. LC and EW drafted the manuscript. All authors critically revised the manuscript and approved the final version.

## Acknowledgements

The authors would like to gratefully acknowledge all COVID-staff of the Public Health Service of Amsterdam. In particular, they would like to thank Anton Janssen for providing population tables, Inge Kramer for linkage with the municipality registration database (BRP) and Anders Boyd for answering statistical questions.

## Availability of data and materials

Information can be obtained from the corresponding author, Liza Coyer.

