## Supplement for "Hospitalisation rates differed by city district and ethnicity during the first wave of COVID-19 in Amsterdam, the Netherlands"

*Contributed equally

**Content** **Page**

Figure 1. COVID-19 notifications from 29 February until 31 May 2020 by notification date, stratified by hospitalisation status, Amsterdam, the Netherlands **2**

Figure 2. Flowchart depicting COVID-19 cases notified in the municipality of Amsterdam between 29 February and 31 May 2020, and linkage of notification data with municipality registration data, the Netherlands **3**

Figure 3. Cumulative hospitalisation rate per 100,000 population in Amsterdam over week of symptom onset between 29 February and 31 May 2020 among those matched with the municipal registration database, by binary representation of city districts **4**

Figure 4. Crude and standardised hospitalisation rates per 100,000 population by migration background and city district in the municipality of Amsterdam between 29 February and 31 May 2020 **5**

Table 1. Characteristics of COVID-19 cases who could and could not be matched to the municipality registration database in Amsterdam, the Netherlands, 29 February to 31 May 2020 **6**

Table 2. Hospitalisation rates by city district among those linked to the registration database, Amsterdam, the Netherlands, 29 February to 31 May 2020 **7**

Table 3. Hospitalisation rates by migration background (first and second generation combined) among those linked to the registration database, Amsterdam, the Netherlands, 29 February to 31 May 2020 **8**

Table 4. Hospitalisation rates by migration history (first and second generation combined) among those aged <60 years linked to the registration database, Amsterdam, the Netherlands, 29 February to 31 May 2020 **9**

Table 5. Hospitalisation rates by migration history (first and second generation combined) among those aged ≥60 years linked to the registration database, Amsterdam, the Netherlands, 29 February to 31 May 2020 **10**

Table 6. Associations with COVID-19 related hospitalization, obtained from a Poisson model with interaction term between migration background / city district, Amsterdam, the Netherlands, 29 February to 31 May 2020 **11**

**Figure S1. COVID-19 notifications from 29 February until 31 May 2020 by notification date, stratified by hospitalisation status, Amsterdam, the Netherlands**

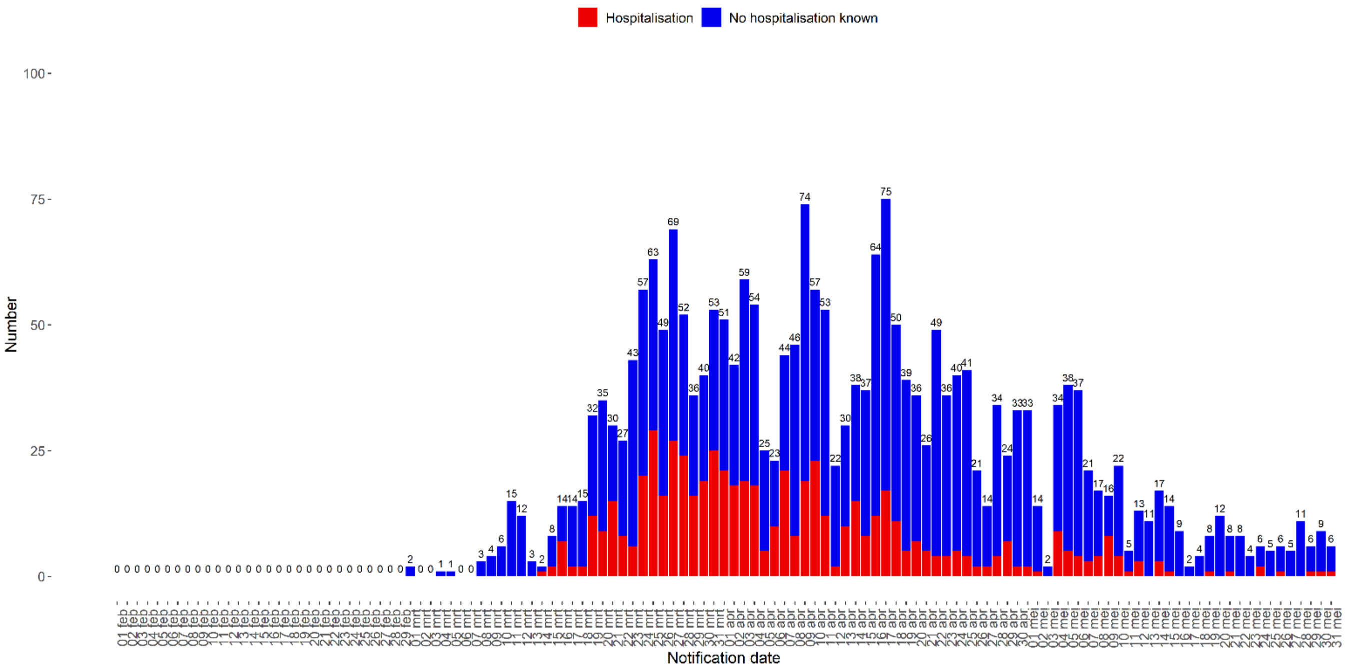

**Figure S2. Flowchart depicting COVID-19 cases notified in the municipality of Amsterdam between 29 February and 31 May 2020, and linkage of notification data with municipality registration data, the Netherlands**

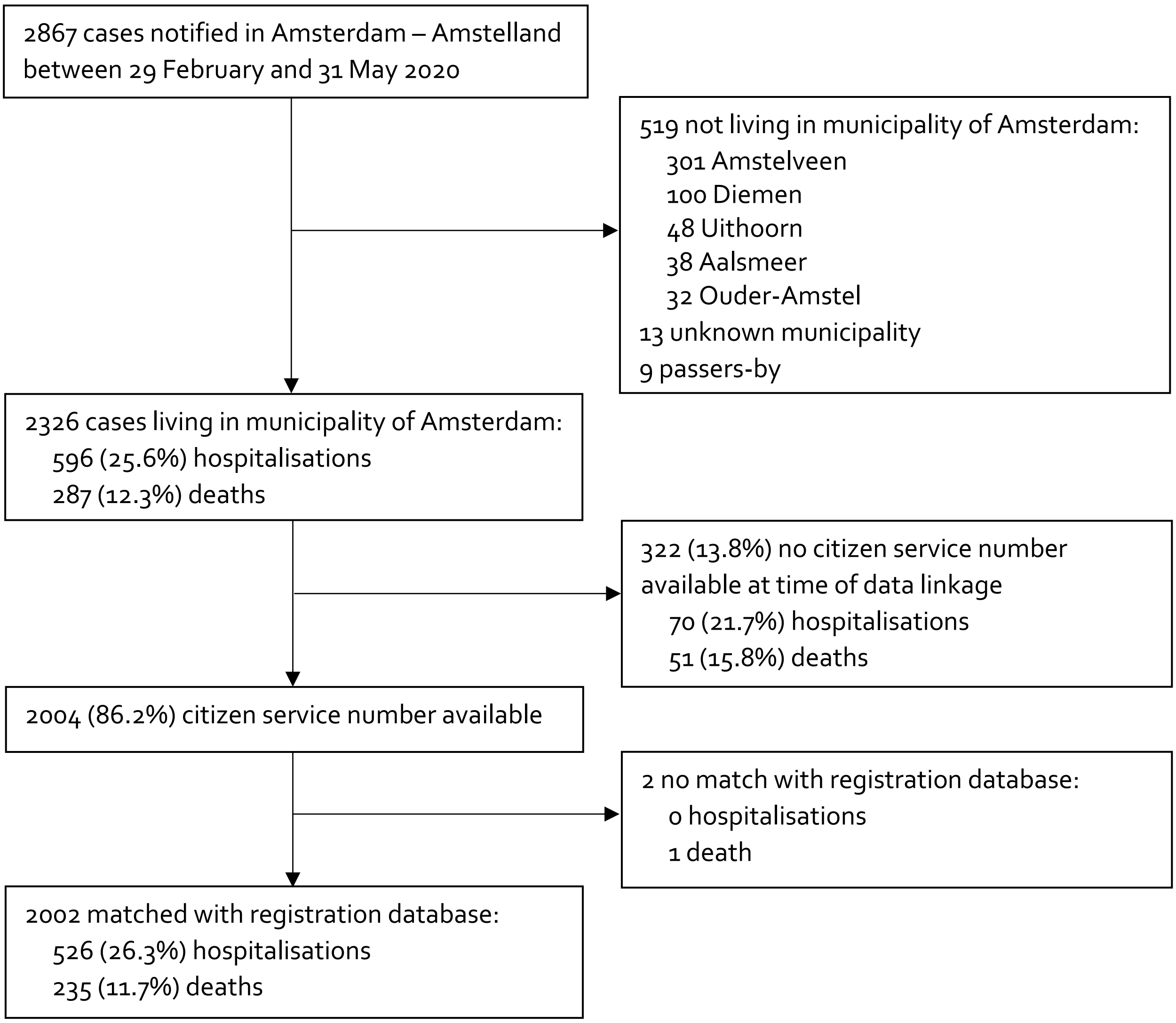

**Figure S3. Cumulative hospitalisation rate per 100,000 population in Amsterdam over week of symptom onset between 29 February and 31 May 2020 among those matched with the municipal registration database, by binary representation of city districts**

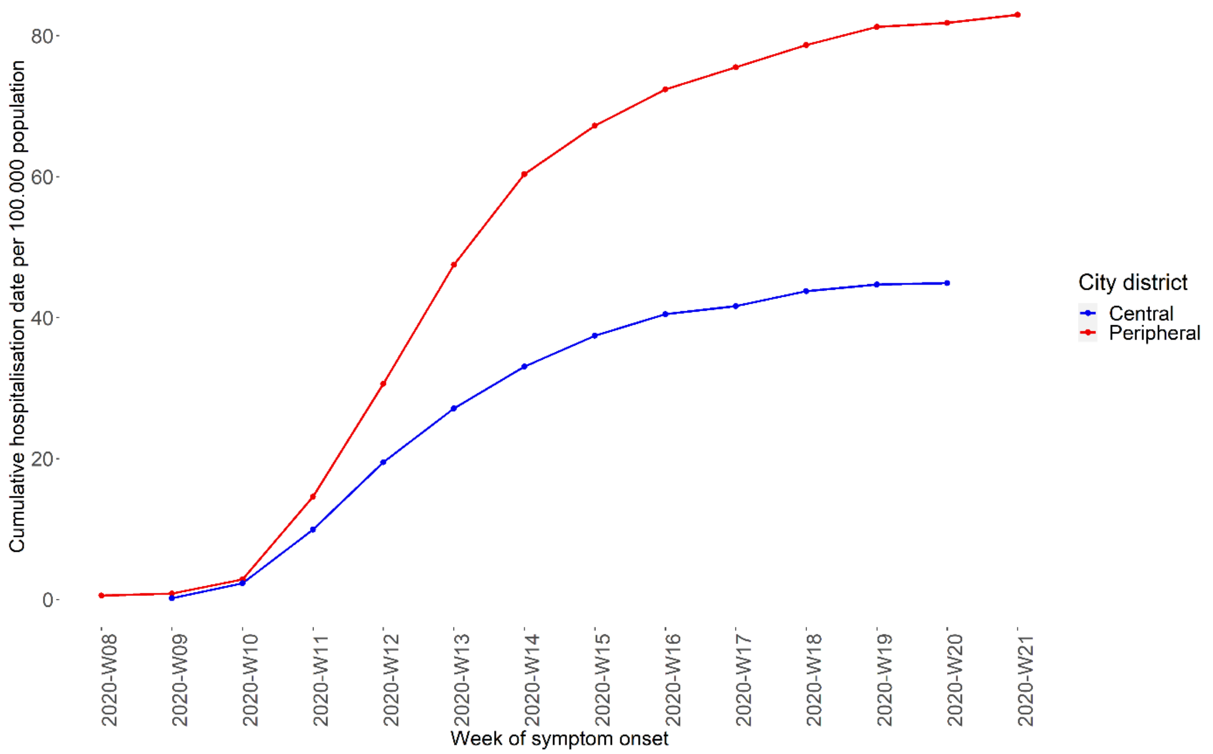

**Figure S4: Crude and standardised hospitalisation rates per 100,000 population by migration background and city district in the municipality of Amsterdam between 29 February and 31 May 2020**

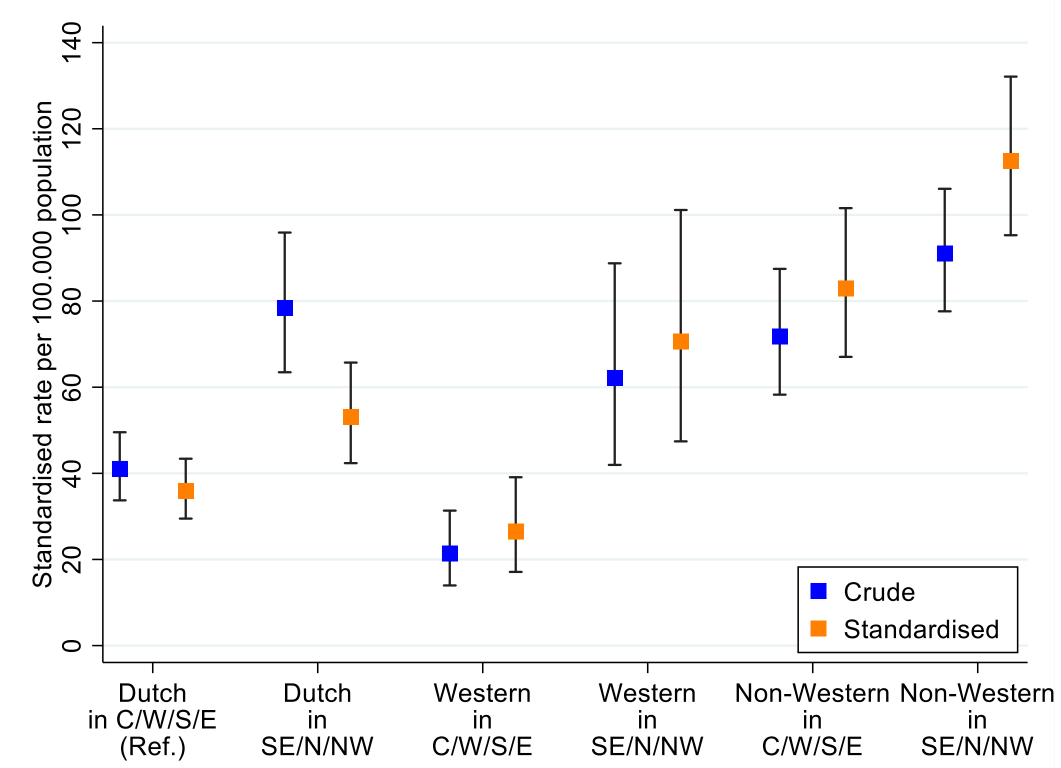

**Table S1. Characteristics of COVID-19 cases who could and could not be matched to the municipality registration database in Amsterdam, the Netherlands, 29 February to 31 May 2020**

| | Total (N=2326) | | Matched (n=2002) | | Not matched (n=324) | |
|---|---|---|---|---|---|---|
| **Characteristic** | **n** | **%** | **n** | **%** | **n** | **%** |
| **Age in years, median [IQR]** | 57 | [37-74] | 56 | [37-73] | 62 | [44-80] |
| **Sex** | | | | | | |
| Female | 1346 | 57.9 | 1181 | 59.0 | 165 | 50.9 |
| Male | 965 | 41.5 | 809 | 40.4 | 156 | 48.2 |
| Unknown | 15 | 0.6 | 12 | 0.6 | 3 | 0.9 |
| **Health care worker (HCW)** | | | | | | |
| No | 1108 | 47.6 | 934 | 46.7 | 174 | 53.7 |
| Yes | 814 | 35.0 | 739 | 46.7 | 75 | 23.2 |
| HCW not in a long-term care facility/unknown location | 500 | 61.4 | 451 | 61.0 | 49 | 65.4 |
| HCW in a long-term care facility | 314 | 38.6 | 288 | 39.0 | 26 | 34.6 |
| Unknown | 404 | 17.3 | 329 | 16.4 | 75 | 23.2 |
| **Resident of a long-term care facility** | | | | | | |
| No/unknown | 1867 | 80.3 | 1673 | 83.6 | 194 | 59.9 |
| Yes | 459 | 19.7 | 329 | 16.4 | 130 | 40.1 |
| **City district** | | | | | | |
| Centre | 162 | 7.0 | 141 | 7.0 | 21 | 6.5 |
| New-West | 433 | 18.6 | 360 | 18.0 | 73 | 22.5 |
| North | 268 | 11.5 | 239 | 12.0 | 29 | 9.0 |
| East | 377 | 16.2 | 330 | 16.5 | 47 | 14.5 |
| West | 341 | 14.7 | 293 | 14.6 | 48 | 14.8 |
| South | 350 | 15.1 | 312 | 15.6 | 38 | 11.7 |
| South-East | 355 | 15.3 | 320 | 16.0 | 45 | 10.8 |
| Unknown | 40 | 1.7 | 7 | 0.4 | 33 | 10.2 |
| **Hospitalised** | | | | | | |
| No/unknown | 1730 | 74.4 | 1476 | 73.7 | 254 | 78.4 |
| Yes | 596 | 25.6 | 526 | 26.3 | 70 | 21.6 |
| **Died** | | | | | | |
| No/unknown | 2039 | 87.7 | 1767 | 88.3 | 272 | 84.0 |
| Yes | 287 | 12.3 | 235 | 11.7 | 52 | 16.1 |

**Abbreviations:** IQR, interquartile range

**Table S2. Hospitalisation rates by city district among those linked to the registration database, Amsterdam, the Netherlands, 29 February to 31 May 2020**

| | Hospital admissions (n) | Population [a] | Crude rate per 100,000 population (95% CI) | Standardised rate per 100,000 population [b] (95% CI) | Standardised rate difference (95% CI) | Standardised rate ratio (95% CI) |
|---|---|---|---|---|---|---|
| **Total** [c] | 523 | 873,055 | 59.90 (54.88-65.27) | | | |
| **City district** | | | | | | |
| Centre | 29 | 872,23 | 33.25 (22.27-47.75) | 26.78 (17.89-38.54) | Ref. | Ref. |
| New-West | 109 | 160,115 | 68.08 (55.90-82.12) | 70.59 (57.93-85.2) | 43.81 (27.29-60.32) | 2.64 (2.22-3.05) |
| North | 79 | 99,705 | 79.23 (62.73-98.75) | 73.12 (57.72-91.36) | 46.34 (27.31-65.37) | 2.73 (2.30-3.16) |
| East | 58 | 142,011 | 40.84 (31.01-52.8) | 44.65 (33.82-57.85) | 17.87 (2.69-33.05) | 1.67 (1.22-2.12) |
| West | 74 | 147,831 | 50.06 (39.31-62.84) | 57.43 (44.99-72.25) | 30.65 (14.20-47.09) | 2.14 (1.71-2.58) |
| South | 72 | 146,471 | 49.16 (38.46-61.9) | 45.65 (35.65-57.58) | 18.86 (4.41-33.31) | 1.70 (1.27-2.14) |
| South-East | 101 | 89,699 | 112.60 (91.71-136.82) | 109.92 (89.32-133.85) | 83.14 (59.35-106.92) | 4.10 (3.69-4.52) |

[a] Population on 1 April 2020
[b] Standardised for age (in 15-year groups) and gender, using the total population of Amsterdam as the standard population
[c] 3 matched cases had missing data

**Table S3. Hospitalisation rates by migration history (first and second generation combined) among those linked to the registration database, Amsterdam, the Netherlands, 29 February to 31 May 2020**

| | Hospital admissions | Population [a] | Crude rate per 100,000 population (95% CI) | Standardised rate per 100,000 population [b] (95% CI) | Standardised rate difference (95% CI) | Standardised rate ratio (95% CI) |
|---|---|---|---|---|---|---|
| **Total [c]** | 523 | 873,055 | 59.90 (54.88-65.27) | | | |
| **Migration background** | | | | | | |
| Netherlands Antilles | 10 | 12,126 | 82.47 (39.55-151.66) | 76.75 (36.79-141.19) | 34.71 (-13.24-82.67) | 1.83 (1.19-2.46) |
| Morocco | 67 | 77,213 | 86.77 (67.25-110.2) | 109.64 (84.08-140.53) | 67.60 (39.71-95.5) | 2.61 (2.32-2.89) |
| Surinam | 78 | 63,944 | 121.98 (96.42-152.24) | 106.61 (83.8-133.71) | 64.57 (39.69-89.46) | 2.54 (2.27-2.8) |
| Turkey | 41 | 44,417 | 92.31 (66.24-125.22) | 129.35 (90.48-179.27) | 87.31 (44.51-130.11) | 3.08 (2.72-3.43) |
| Ghana | 17 | 12,883 | 131.96 (76.87-211.28) | 178.79 (52.99-438.28) | 136.75 (-29.83-303.33) | 4.25 (3.31-5.19) |
| Other non-Western | 49 | 106,137 | 46.17 (34.15-61.03) | 67.34 (47.20-93.15) | 25.3 (2.58-48.01) | 1.60 (1.25-1.96) |
| Total non-Western | 262 | 316,720 | 82.72 (73.01-93.37) | 99.08 (87.08-112.28) | 57.05 (43.34-70.75) | 2.36 (2.17-2.54) |
| Total Western | 57 | 169,814 | 33.57 (25.42-43.49) | 41.06 (30.96-53.4) | -0.98 (-13.28-11.32) | 0.98 (0.68-1.27) |
| Dutch | 204 | 386,521 | 52.78 (45.78-60.54) | 42.04 (36.38-48.32) | Ref. | Ref. |

[a] Population on 1 April 2020
[b] Standardised for age (in 15-year groups) and gender, using the total population of Amsterdam as the standard population
[c] 3 matched cases had missing data

**Table S4. Hospitalisation rates by migration history (first and second generation combined) among those aged <60 years linked to the registration database, Amsterdam, the Netherlands, 29 February to 31 May 2020**

| | Hospital admissions | Population [a] | Crude rate per 100,000 population (95% CI) | Standardised rate per 100,000 population [b] (95% CI) | Standardised rate difference (95% CI) | Standardised rate ratio (95% CI) |
|---|---|---|---|---|---|---|
| **Total** | 197 | 715,610 | 27.53 (23.82-31.65) | | | |
| **Migration background** | | | | | | |
| Netherlands Antilles | 3 | 10,044 | 29.87 (6.16-87.29) | 28.83 (5.94-84.25) | 13.64 (-19.27-46.55) | 1.90 (0.73-3.07) |
| Morocco | 36 | 68,124 | 52.84 (37.01-73.16) | 59.92 (41.94-83) | 44.74 (24.65-64.82) | 3.95 (3.51-4.38) |
| Surinam | 34 | 49,825 | 68.24 (47.26-95.36) | 59.79 (40.98-84.26) | 44.6 (23.53-65.67) | 3.94 (3.49-4.39) |
| Turkey | 19 | 39,898 | 47.62 (28.67-74.37) | 47 (28.28-73.42) | 31.81 (10.22-53.4) | 3.09 (2.56-3.63) |
| Ghana | 7 | 11,056 | 63.31 (25.46-130.45) | 48.14 (18.95-100.46) | 32.95 (-3.64-69.54) | 3.17 (2.36-3.98) |
| Other non-Western | 30 | 96,978 | 30.93 (20.87-44.16) | 33.38 (22.39-47.89) | 18.2 (5.32-31.07) | 2.20 (1.74-2.66) |
| Total non-Western | 129 | 275,925 | 46.75 (39.03-55.55) | 48.3 (40.32-57.39) | 33.11 (23.71-42.52) | 3.18 (2.85-3.52) |
| Total Western | 21 | 146,910 | 14.29 (8.85-21.85) | 14.83 (9.12-22.79) | -0.36 (-8.12-7.40) | 0.98 (0.46-1.5) |
| Dutch | 47 | 292,775 | 16.05 (11.8-21.35) | 15.19 (11.14-20.22) | Ref. | Ref. |

[a] Population on 1 April 2020
[b] Standardised for age (in 15-year groups) and gender, using the total population of Amsterdam as the standard population

**Table S5. Hospitalisation rates by migration history (first and second generation combined) among those aged ≥60 years linked to the registration database, Amsterdam, the Netherlands, 29 February to 31 May 2020**

| | Hospital admissions | Population [a] | Crude rate per 100,000 population (95% CI) | Standardised rate per 100,000 population [b] (95% CI) | Standardised rate difference (95% CI) | Standardised rate ratio (95% CI) |
|---|---|---|---|---|---|---|
| **Total** | 326 | 157,445 | 207.06 (185.19-230.8) | | | |
| **Migration background** | | | | | | |
| Netherlands Antilles | 7 | 2,082 | 336.22 (135.18-692.73) | 221.84 (89.13-457.24) | 34.03 (-133.26-201.32) | 1.18 (0.42-1.94) |
| Morocco | 31 | 9,089 | 341.07 (231.74-484.12) | 369.25 (235.99-550.51) | 181.44 (29.8-333.09) | 1.97 (1.53-2.4) |
| Surinam | 44 | 14,119 | 311.64 (226.44-418.36) | 309.29 (212.07-435.76) | 121.49 (10.58-232.39) | 1.65 (1.27-2.03) |
| Turkey | 22 | 4,519 | 486.83 (305.1-737.07) | 537.6 (306.7-874.28) | 349.8 (83.8-615.8) | 2.86 (2.34-3.38) |
| Ghana | 10 | 1,827 | 547.35 (262.47-1006.59) | 993.63 (94.21-3901.41) | 805.83 (-675.28-2286.93) | 5.29 (3.79-6.79) |
| Other non-Western | 19 | 9,159 | 207.45 (124.9-323.95) | 256.7 (127.5-460.89) | 68.89 (-86.81-224.59) | 1.37 (0.75-1.98) |
| Total non-Western | 133 | 40,795 | 326.02 (272.97-386.37) | 344.08 (278.02-421.12) | 156.27 (80.16-232.39) | 1.83 (1.57-2.09) |
| Total Western | 36 | 22,904 | 157.18 (110.09-217.6) | 180.69 (122.95-256.16) | -7.12 (-77.59-63.35) | 0.96 (0.57-1.35) |
| Dutch | 157 | 93,746 | 167.47 (142.3-195.82) | 187.81 (158.26-221.27) | Ref. | Ref. |

[a] Population on 1 April 2020
[b] Standardised for age (in 15-year groups) and gender, using the total population of Amsterdam as the standard population

**Table S6. Associations with COVID-19 related hospitalization, obtained from a Poisson model with interaction term between migration background / city district, Amsterdam, the Netherlands, 29 February to 31 May 2020**

| Factor | RR (95% CI) | *P*-value |
|---|---|---|
| **Migration background** | | |
| None (ethnic-Dutch) | 1 | |
| Western | 0.74 (0.47-1.11) | 0.16 |
| Non-Western | 2.33 (1.77-3.06) | <0.001 |
| **City district** | | |
| Central (C/W/S/E) | 1 | |
| Peripheral (SE/N/NW) | 1.49 (1.13-1.96) | 0.005 |
| **Migration * district term** | | |
| Peripheral/ethnic-Dutch | 1 | |
| Peripheral/Western | 1.78 (0.99-3.24) | 0.06 |
| Peripheral/Non-Western | 0.92 (0.63-1.34) | 0.66 |
| **Sex** | | |
| Male | 1 | |
| Female | 0.59 (0.49-0.7) | <0.001 |
| **Age** | | |
| <45 years | 1 | |
| 45-59 years | 4.96 (3.74-6.61) | <0.001 |
| 60-74 years | 13.49 (10.45-17.59) | <0.001 |
| ≥75 years | 23.63 (17.75-31.62) | <0.001 |

Likelihood ratio test *P*-value this model vs. model without interaction term = 0.08
